# CerebAI: Explainable Three-Class Stroke CT Classification via ConvNeXt and Integrated Gradients

**DOI:** 10.64898/2026.07.03.26357233

**Authors:** Adarsh R Shenoy, Tanya Mendez

**Affiliations:** Independent Researcher; Department of Robotics and Artificial Intelligence, NMAM Institute of Technology (NMAMIT), Nitte (Deemed to be University), Udupi, India

**Keywords:** Stroke classification, computed tomography, ConvNeXt, integrated gradients, explainable AI, medical imaging, deep learning

## Abstract

Stroke is a leading cause of death and long-term disability worldwide, affecting approximately 15 million individuals annually. Prompt and accurate subtype differentiation between ischemic and hemorrhagic stroke is clinically critical, as the two conditions demand diametrically opposite interventions—thrombolytic therapy versus surgical decompression. Yet the majority of existing deep learning approaches reduce this problem to binary detection, and virtually none address the opacity of their decision-making in a clinically actionable manner. We present CerebAI, an explainable, deployment-oriented three-class CT stroke classification system built on a fine-tuned ConvNeXt-Base backbone with Integrated Gradients (IG) attribution. Trained on 6,774 non-contrast CT scans stratified across No Stroke, Ischemic Stroke, and Hemorrhagic Stroke, CerebAI achieves a weighted F1-score of 0.9746 (95% CI: [0.9625, 0.9851]), accuracy of 97.47%, macro-averaged AUC of 0.9921, mean Intersection-over-Union (mIoU) of 0.9276, Expected Calibration Error (ECE) of 0.0115, mean Brier Score of 0.0150, and Cohen’s *κ* of 0.9483—surpassing ResNet-50, EfficientNet-B4, and Vision Transformer (ViT-B/16) baselines across all reported metrics. Integrated Gradients produce pixel-precise saliency maps that localize pathological regions with greater anatomical fidelity than Gradient-weighted Class Activation Mapping (Grad-CAM), a finding we support with side-by-side qualitative comparison. CerebAI additionally incorporates a native DICOM processing pipeline to facilitate future clinical translation. Code and model weights are publicly available to support reproducibility and further research.

## I. Introduction

Stroke ranks among the most devastating neurological emergencies globally. According to the Global Burden of Disease Study 2019, approximately 12.2 million new stroke events occur each year, resulting in 6.55 million deaths and making stroke the second leading cause of mortality worldwide [1]. The World Health Organization estimates that one in four adults over the age of 25 will experience a stroke during their lifetime, and survivors frequently endure lasting motor, cognitive, and communicative impairments [2].

Central to acute stroke management is the principle that *time is brain*: each minute of delayed treatment results in the loss of approximately 1.9 million neurons [3]. Non-contrast computed tomography (NCCT) remains the gold-standard first-line imaging modality in emergency settings worldwide due to its rapid acquisition speed, widespread availability, and proven sensitivity for hemorrhagic events [4]. However, accurate radiological interpretation of NCCT requires experienced neuroradiologists who may not be immediately available in resource-limited or rural settings. This diagnostic bottleneck motivates the development of reliable, automated, and clinically transparent AI-assisted tools for acute stroke triage.

A fundamental clinical distinction separates the two principal stroke subtypes. Ischemic stroke, caused by thromboembolic occlusion of cerebral vasculature, constitutes approximately 87% of all strokes and is treated with intravenous tissue plasminogen activator (tPA) within a narrow therapeutic window [5]. Hemorrhagic stroke, caused by rupture of a blood vessel with resultant intracranial hemorrhage, may require urgent surgical evacuation [6]. Critically, administering tPA to a hemorrhagic patient can be life-threatening. The clinical imperative for accurate, rapid subtype classification is therefore absolute.

Despite the proliferation of deep learning approaches to stroke imaging, three significant gaps persist in the literature. **First**, the majority of models address binary detection—stroke versus no stroke—rather than the clinically necessary three-class subtype problem. **Second**, nearly all existing models are black boxes, providing classification decisions without indicating which image regions drove them, rendering them unsuitable for clinical trust and regulatory scrutiny. **Third**, existing research prototypes rarely provide a DICOM-compatible inference pipeline, creating friction for real-world hospital integration.

We address all three gaps with **CerebAI**, making the following contributions:

- A fine-tuned **ConvNeXt-Base** model for three-class NCCT stroke classification (No Stroke, Ischemic, Hemorrhagic) achieving superior performance across seven evaluation metrics compared to ResNet-50, EfficientNet-B4, and ViT-B/16.
- Pixel-level clinical explainability via **Integrated Gradients** (IG), grounded in axiomatic attribution theory, producing anatomically meaningful saliency maps that outperform Grad-CAM in spatial precision.
- A **DICOM-native processing pipeline** enabling direct inference from clinical DICOM files to facilitate future clinical integration.
- **Comprehensive evaluation** including per-class metrics, bootstrap confidence intervals, calibration analysis (ECE, Brier Score), IoU per class, three-baseline comparison, and ablation study.

## II. Related Work

### A. Deep Learning for Stroke CT Analysis

The application of convolutional neural networks to brain CT analysis has accelerated considerably over the past decade. Kuo et al. [7] demonstrated that ResNet-based architectures could detect acute ischemic changes in NCCT with radiologist-level sensitivity. Subsequent studies expanded to hemorrhage detection: Chang et al. [9] proposed a hybrid 3D CNN approach for hemorrhage subtype classification, while Arbabshirani et al. [10] deployed a deep learning triage system for critical radiology findings including intracranial hemorrhage. However, these approaches predominantly address binary classification or hemorrhage-only detection, and very few simultaneously tackle all three clinically relevant categories: normal, ischemic, and hemorrhagic. CerebAI explicitly frames this as a three-class problem, reflecting real emergency department triage requirements.

### B. Explainability in Medical AI

The opacity of deep neural networks poses a fundamental obstacle to clinical adoption. Selvaraju et al. [11] introduced Gradient-weighted Class Activation Mapping (Grad-CAM), which computes saliency via gradients of the classification score with respect to the final convolutional feature maps. While widely adopted, Grad-CAM is constrained to the spatial resolution of the terminal feature layer (7×7 for ConvNeXt-Base), yielding coarse, block-like attributions.

Sundararajan et al. [12] proposed Integrated Gradients (IG), grounded in two mathematical axioms—*sensitivity* and *implementation invariance*—that gradient-based methods such as Grad-CAM fundamentally violate. IG computes the integral of gradients along a straight path from a reference baseline to the input, yielding pixel-level attributions without architectural constraints. In medical imaging, where a clinician must understand which specific pixels drove a decision, pixel-level precision is essential.

### C. ConvNeXt and Modern CNN Architectures

Liu et al. [13] introduced ConvNeXt as a pure convolutional architecture retaining the scalability and training recipes of Vision Transformers while preserving spatial inductive biases suited to image data. ConvNeXt-Base employs 7×7 depth-wise convolutions, inverted bottleneck blocks, GELU activations [14], and layer normalization, outperforming comparably scaled ViT models on ImageNet. In medical imaging, these convolutional inductive biases remain advantageous when data volume is limited—as our ablation demonstrates, with F1 collapsing from 0.9746 to 0.5403 when removing ImageNet pretraining.

## III. Dataset and Preprocessing

### A. Dataset

We utilize a publicly available non-contrast CT stroke dataset comprising **6**,**774 axial brain CT slices** across three diagnostic classes: No Stroke (4,551), Ischemic Stroke (1,130), and Hemorrhagic Stroke (1,093) [28]. Images were acquired at an average resolution of 514×512 pixels and stored as PNG exports from DICOM sources. The scan provenance and original acquisition institution are not documented in the publicly released dataset; future work should validate findings on datasets with confirmed IRB approval and known scanner demographics.

The dataset exhibits a natural class imbalance reflecting real-world stroke epidemiology: No Stroke cases constitute 67.2% of the dataset, while Ischemic and Hemorrhagic each constitute approximately 16.4%. A stratified random split was applied: 4,741 images for training (70%), 1,362 for validation (20%), and 671 for testing (10%), with class proportions preserved across all partitions using fixed seed *s* = 42. Table I summarizes the statistics.

**TABLE I.** Dataset Statistics and Split Distribution.

| Class | Total | % | Train | Val | Test |
| --- | --- | --- | --- | --- | --- |
| No Stroke | 4,551 | 67.2 | 3,185 | 910 | 451 |
| Ischemic | 1,130 | 16.7 | 791 | 226 | 112 |
| Hemorrhagic | 1,093 | 16.1 | 765 | 220 | 108 |
| **Total** | **6,774** | **100** | **4,741** | **1,362** | **671** |

### B. Preprocessing and Augmentation

All images were resized to 224×224 pixels and converted from grayscale to three-channel RGB to match ImageNet pretraining expectations, normalized to [−1, 1] with *µ* = 0.5, *σ* = 0.5.

During training, a stochastic augmentation pipeline was applied via Albumentations [20]: horizontal and vertical flipping (*p* = 0.5), rotation (±20^*◦*^, *p* = 0.7), shift-scale-rotate (*p* = 0.7), elastic transform (*p* = 0.5), grid distortion (*p* = 0.5), optical distortion (*p* = 0.5), brightness-contrast perturbation (*p* = 0.7), Gaussian noise (*p* = 0.5), and coarse dropout (*p* = 0.5). Validation and test images received only resize and normalize. Ablation confirms this pipeline contributes +0.15% F1 over no-augmentation training.

## IV. Methodology

### A. System Architecture

Fig. 1 illustrates the end-to-end CerebAI pipeline. A CT scan (PNG or DICOM) is preprocessed into a 224×224×3 tensor, passed through a fine-tuned ConvNeXt-Base backbone, and classified into one of three stroke categories. In parallel, Integrated Gradients generates a pixel-level attribution map identifying the regions most responsible for the prediction.

**Fig. 1.**
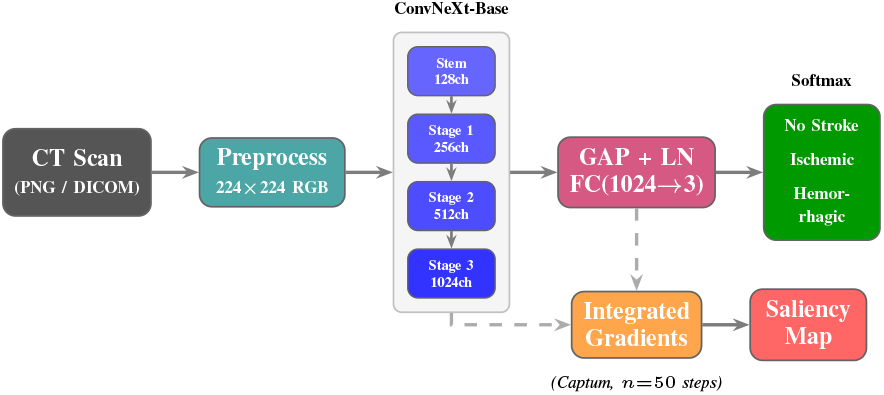
CerebAI end-to-end pipeline. A CT image (PNG or DICOM) is preprocessed and classified by a fine-tuned ConvNeXt-Base. In parallel, Integrated Gradients generates pixel-level attribution maps. Dashed arrows indicate the gradient flow path for attribution.

### B. ConvNeXt-Base Backbone

ConvNeXt-Base [13] is a hierarchical CNN comprising a patchify stem followed by four progressive stages containing 3, 3, 27, and 3 ConvNeXt blocks, yielding channel widths of 128, 256, 512, and 1024. Each block employs a 7 × 7 depth-wise convolution, inverted bottleneck structure (4 × channel expansion), GELU activation [14], and Layer Normalization. The classifier head consists of Global Average Pooling, Layer Normalization, and a linear projection from 1,024 to 3 logits.

We initialize with ImageNet-1k pretrained weights via `timm` [21] and fine-tune all 87.57 million parameters jointly. The model requires 15.35 GFLOPs per forward pass and achieves 17.04 ± 5.64 ms inference latency per image on an NVIDIA Tesla T4 GPU.

### C. Training Procedure

Training was conducted for 50 epochs with batch size 16, AdamW optimizer [18] (lr = 10^−4^, weight decay = 10^−5^), and CrossEntropyLoss. A ReduceLROnPlateau scheduler (factor 0.5, patience 5) monitored validation weighted F1. Mixed-precision training via PyTorch AMP [19] with gradient scaling was used throughout. All experiments used seed 42 with PyTorch deterministic mode for full reproducibility. Best checkpoint saved at epoch 50 (Val F1 = 0.9718).

### D. Integrated Gradients Attribution

Let *f*_*c*_(**x**) denote the *c*-th logit for input **x** ∈ ℝ^*H×W×*3^ and baseline **x**^*′*^ = **0**. Integrated Gradients [12] defines the attribution of pixel *i* for class *c* as:

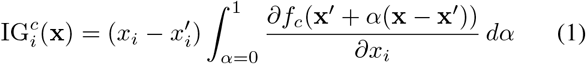

The integral in (1) is approximated via Riemann sum with *n* = 50 steps using Captum [22]. A zero-tensor baseline represents the absence of imaging signal— a principled choice for CT where black corresponds to air or background. IG satisfies the *Completeness* axiom: 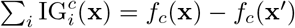, ensuring attributions fully account for the prediction gap—a property Grad-CAM does not satisfy.

## V. Experiments and Results

### A. Quantitative Performance

Table II reports per-class metrics on the held-out test set (*n* = 671). CerebAI achieves near-perfect classification of No Stroke (F1=0.9868, recall=0.9933), Hemorrhagic Stroke F1=0.9619, and Ischemic Stroke F1=0.9375— reflecting the inherent subtlety of early-stage ischemic changes on NCCT.

**TABLE II.** Per-Class Performance on Test Set (*n* = 671)

| Class | Prec. | Recall | F1 | AUC |
| --- | --- | --- | --- | --- |
| No Stroke | 0.9803 | 0.9933 | 0.9868 | 0.9890 |
| Ischemic Stroke | 0.9375 | 0.9375 | 0.9375 | 0.9933 |
| Hemorrhagic | 0.9902 | 0.9352 | 0.9619 | 0.9975 |
| Weighted Avg | 0.9748 | 0.9747 | 0.9746 | — |
| Accuracy: 0.9747 Macro AUC: 0.9921 |  |  |  |  |
| Cohen's $\kappa$: 0.9483 mIoU: 0.9276 | | | | |
| ECE: 0.0115 Mean Brier: 0.0150 |  |  |  |  |
| F1 95% CI: [0.9625, 0.9851] (bootstrap, $n=1000$) | | | | |

Cohen’s *κ* = 0.9483 indicates *almost perfect* agreement beyond chance [23]. The ECE of 0.0115 and mean Brier Score of 0.0150 demonstrate well-aligned confidence scores—a critical property for clinical risk communication. The bootstrap 95% CI [0.9625, 0.9851] confirms statistical robustness.

### B. Training Dynamics

Fig. 2 presents training curves over 50 epochs. Training loss decreases monotonically from 0.597 to 0.026; validation loss stabilizes in [0.10, 0.18] from epoch 20 onward, indicating effective regularization with minimal overfitting. Validation F1 rises rapidly in the first 15 epochs (0.855 → 0.962), then steadily to 0.9718 at epoch 50.

**Fig. 2.**
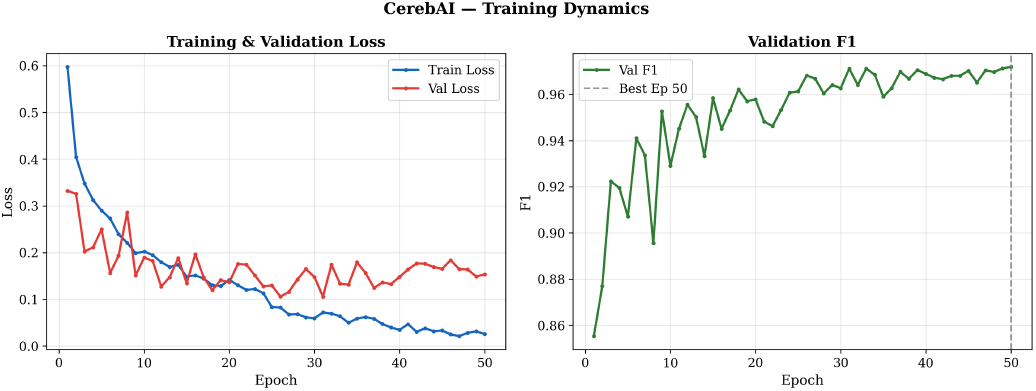
Training dynamics over 50 epochs. *Left*: Training and validation cross-entropy loss. *Right*: Weighted validation F1 trend. Dashed line marks best checkpoint (epoch 50).

### C. Confusion Matrix Analysis

Fig. 3 shows the test set confusion matrix. Of 451 No Stroke cases, 448 are correctly classified (99.3%). Of 112 Ischemic cases, 105 are correct (93.8%). Of 108 Hemorrhagic cases, 101 are correct (93.5%). Critically, zero No Stroke cases are misclassified as Hemorrhagic and vice versa—the two highest-stakes error types are absent.

**Fig. 3.**
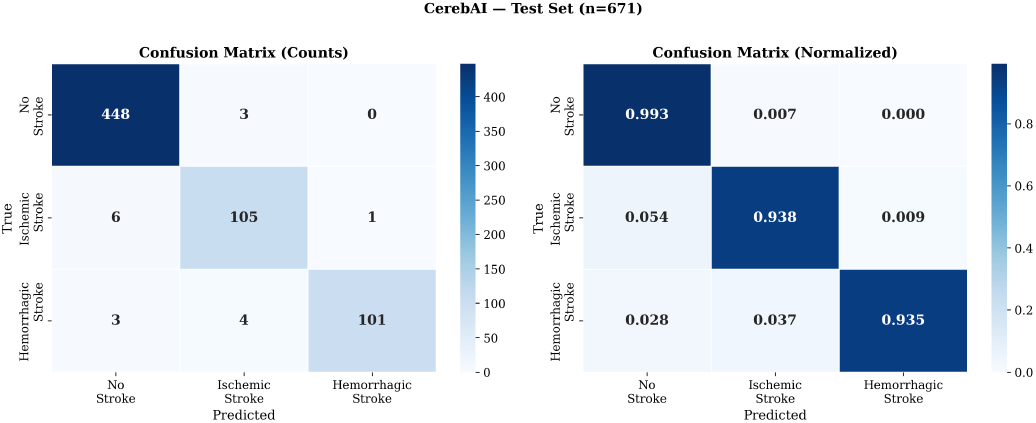
Confusion matrices (*n* = 671). *Left*: Absolute counts. *Right*: Row-normalized proportions. No cross-class confusions exist between No Stroke and Hemorrhagic categories.

### D. ROC and Calibration Analysis

Fig. 4 presents per-class ROC curves using one-vs-rest decomposition. All three classes achieve AUC *>* 0.98, with Hemorrhagic Stroke reaching AUC=0.9975. Macro-averaged AUC of 0.9921 confirms consistent discriminative power across all classes.

**Fig. 4.**
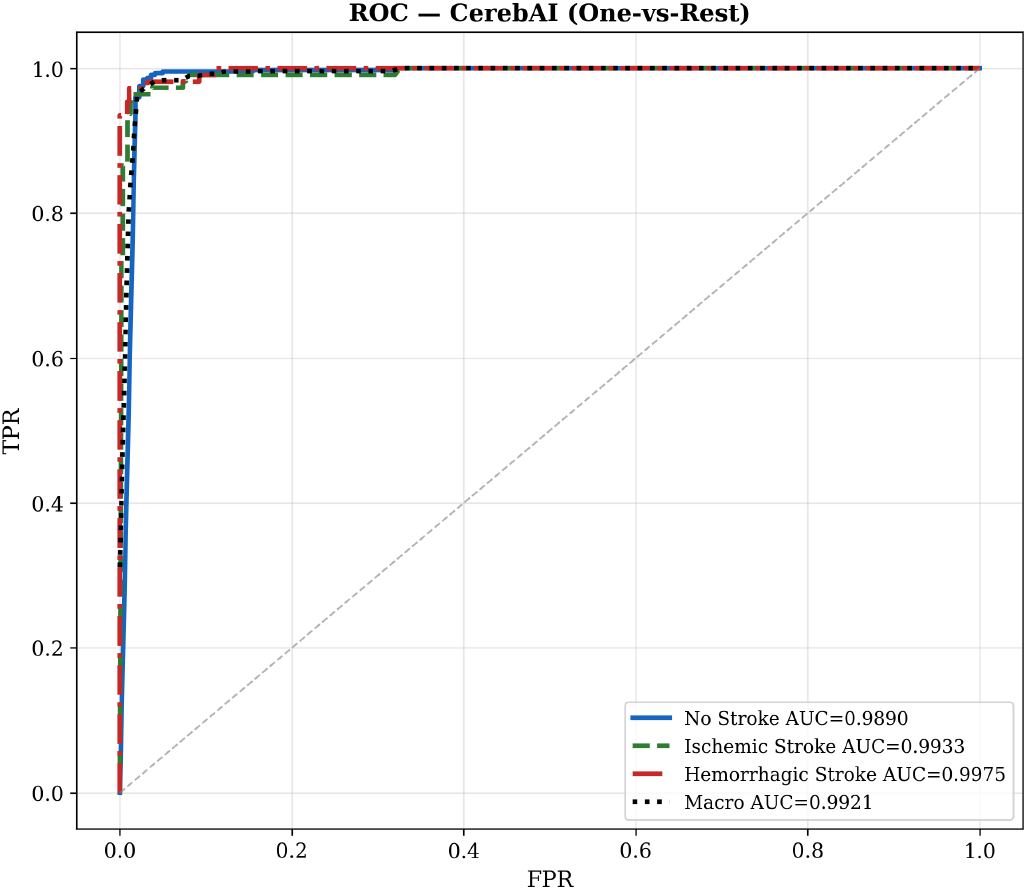
Multiclass ROC curves (one-vs-rest). All per-class AUC values exceed 0.989. Macro-averaged AUC=0.9921.

Fig. 5 presents the quantile-binned reliability diagram. All class-wise curves track closely along the perfect calibration diagonal. The ECE of 0.0115 is substantially lower than typical uncalibrated deep networks (*>* 0.05) [24].

**Fig. 5.**
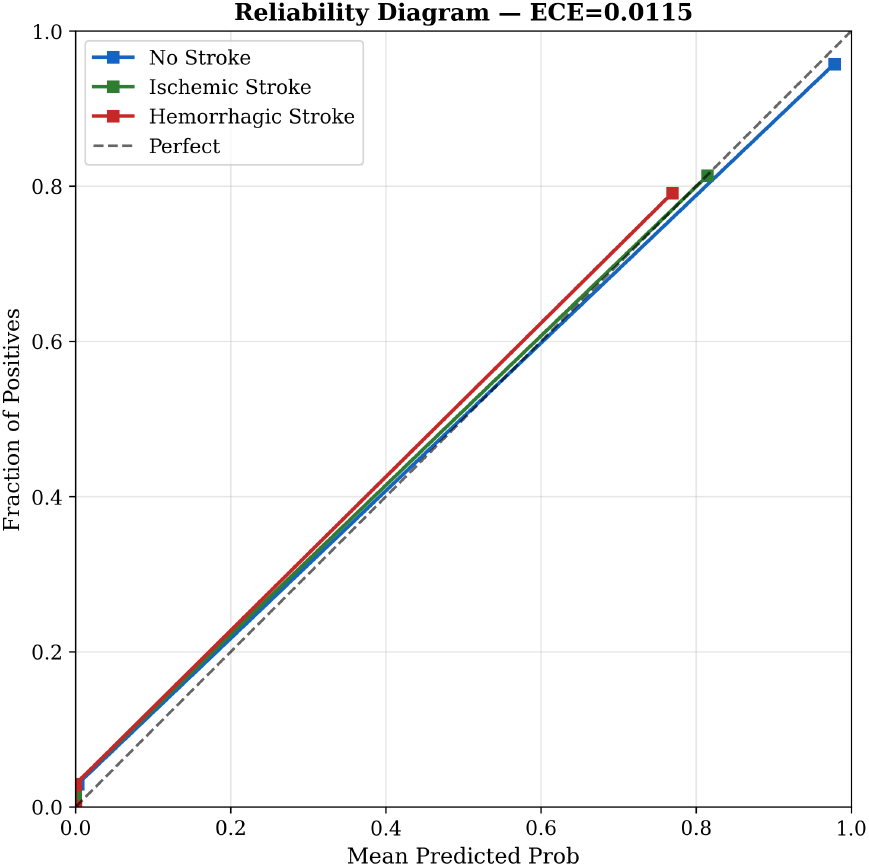
Quantile-binned reliability diagram. All class curves approximate the diagonal. ECE=0.0115; Mean Brier Score=0.0150.

### E. Comparison with Baseline Models

Table III compares CerebAI against three established backbones trained under identical conditions: same data, same split, same optimizer, same augmentation, AdamW *η* = 10^−4^, maximum 30 epochs, early stopping (patience=7).

**TABLE III.** Comparison with Baseline Models (Test Set, *n* = 671)

| Model | Acc. | F1 (W) | AUC | $\kappa$ | Param. (M) | Inf. (ms) |
| --- | --- | --- | --- | --- | --- | --- |
| ResNet-50 [15] | 0.9255 | 0.9238 | 0.9829 | 0.8455 | 23.5 | 6.28 |
| ViT-B/16 [17] | 0.9046 | 0.9025 | 0.9669 | 0.7994 | 85.8 | 21.50 |
| EffNet-B4 [16] | 0.9613 | 0.9608 | 0.9911 | 0.9200 | 17.6 | 15.45 |
| **CerebAI** | **0.9747** | **0.9746** | **0.9932** | **0.9483** | 87.6 | 17.04 |

CerebAI outperforms all baselines across every metric. Relative to the strongest baseline (EfficientNet-B4), CerebAI achieves +1.38% weighted F1, +0.21% macro AUC, and +2.83 points in Cohen’s *κ*. ViT-B/16 underperforms despite a comparable parameter count (85.8M vs. 87.6M), supporting the view that convolutional inductive biases remain advantageous at moderate dataset scales.

### F. Ablation Study

Table IV isolates two key design choices.

**TABLE IV.** Ablation Study — Convnext-Base Variants.

| Configuration | Accuracy | F1 (W) |
| --- | --- | --- |
| w/o ImageNet Pretraining | 0.6721 | 0.5403 |
| w/o Data Augmentation | 0.9732 | 0.9731 |
| **CerebAI (Full)** | **0.9747** | **0.9746** |

#### Pretraining

Removing ImageNet-1k pretraining causes catastrophic F1 collapse from 0.9746 to 0.5403 (− 43.43%), with early stopping at epoch 7. Transfer learning is not merely beneficial but essential at this dataset scale.

#### Augmentation

Training without stochastic augmentation yields F1=0.9731 vs. 0.9746 (+0.15%). While marginal in aggregate, augmentation reduces overfitting risk and improves generalization to unseen scanner variability.

### G. Explainability Analysis

#### 1) Multi-Class Integrated Gradients

Fig. 6 presents IG attribution maps for two correctly predicted examples per class (six total). For No Stroke cases, attributions distribute across the normal sulcal-gyral pattern and ventricular anatomy. For Ischemic Stroke, attributions highlight regions of subtle hypodensity and loss of gray-white differentiation— early CT signs of cerebral infarction. For Hemorrhagic Stroke, attributions concentrate precisely on the hyperdense intraparenchymal hematoma, the canonical CT signature of intracerebral hemorrhage.

**Fig. 6.**
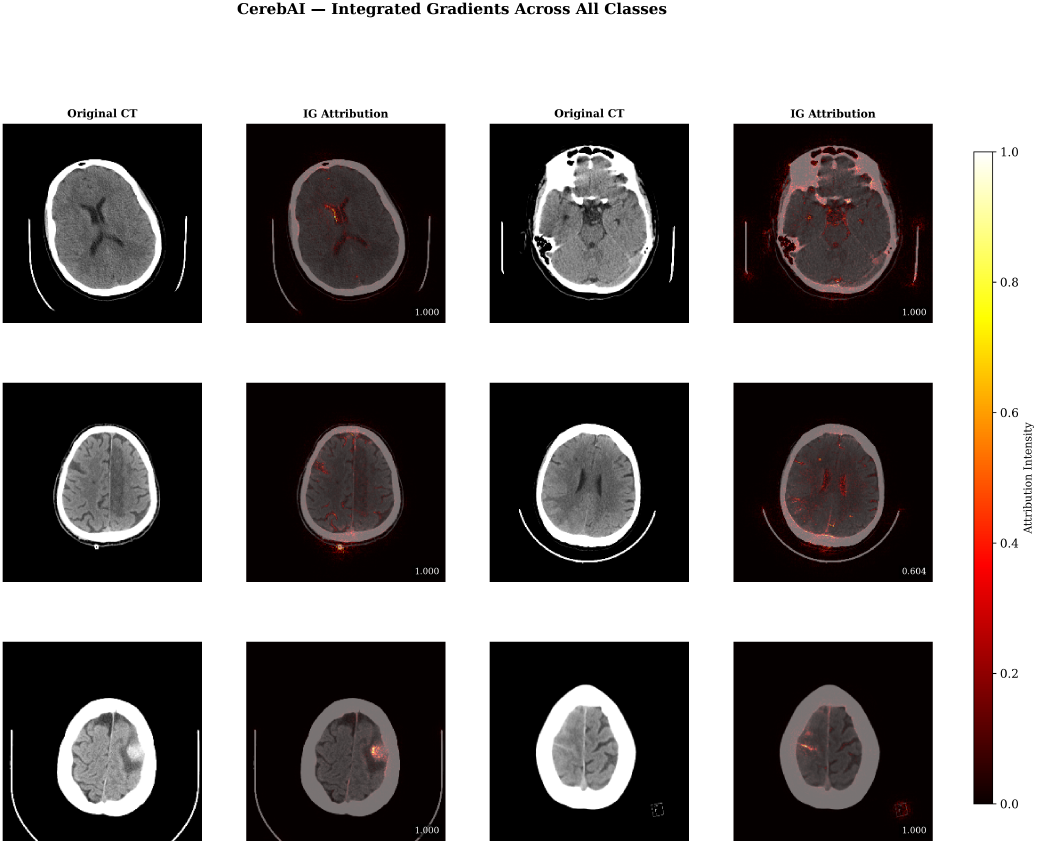
Integrated Gradients attribution maps. Rows: No Stroke, Ischemic Stroke, Hemorrhagic Stroke. Each pair shows the original CT and its IG overlay. Hotter colors indicate higher attribution intensity. Confidence scores shown in lower-right corner of each IG map.

#### 2) Grad-CAM vs. Integrated Gradients

Fig. 7 directly compares Grad-CAM and IG on an identical Hemorrhagic Stroke CT (confidence = 1.000 for both). Grad-CAM produces a coarse, spatially diffuse activation map constrained to 7×7 spatial resolution, requiring bilinear upsampling that introduces significant blur. IG produces a pixel-precise map with concentrated attribution directly on the hyperdense hemorrhagic lesion. In a clinical context where a radiologist must verify which specific region drove the model’s decision, pixel-level precision is not optional.

**Fig. 7.**
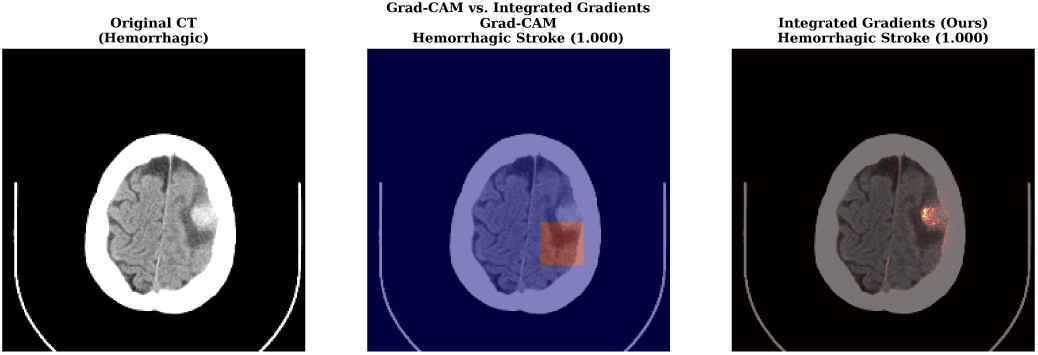
Explainability comparison on a Hemorrhagic Stroke CT. *Left*: Original CT. *Center*: Grad-CAM—diffuse, coarse activation. *Right*: CerebAI IG—precise localization of the hyperdense hematoma. Both methods predict Hemorrhagic Stroke (confidence = 1.000).

## VI. Discussion

### A. Clinical Implications

CerebAI’s Cohen’s *κ* of 0.9483 situates the model in the *almost perfect* agreement category [23], comparable to inter-radiologist agreement for stroke CT in emergency settings [26]. The ECE of 0.0115 ensures that CerebAI’s stated confidence levels closely reflect empirical accuracy—a necessary property for any system informing triage urgency decisions.

The asymmetric error pattern in the confusion matrix is clinically favorable: zero cross-class confusions between No Stroke and Hemorrhagic—the highest-stakes error pair. The six ischemic-to-normal misclassifications warrant attention in future work, as these represent potential missed diagnoses. Pixel-level IG attribution provides an auditable decision trail: a clinician can verify that the model attended to the correct anatomical region before acting on its recommendation—addressing a core requirement for AI medical device regulatory pathways.

### B. Limitations

We acknowledge four key limitations. **(1) Single-source dataset:** All images originate from one publicly available source without documented institutional provenance, scanner specifications, or IRB approval. External validation on established benchmarks such as RSNA Intracranial Hemorrhage [27] or PhysioNet CQ500 is required before any clinical application. **(2) Slice-level classification:** CerebAI classifies individual axial slices rather than full volumetric studies; clinical diagnosis integrates findings across multiple slices. **(3) No cross-validation:** Results are reported on a single stratified test split; k-fold cross-validation would strengthen statistical claims. **(4) No prospective validation:** All results are retrospective on held-out data; no clinical trial has been conducted.

### C. Future Work

Planned extensions include multi-center external validation, volumetric 3D classification via ConvNeXt-3D, PACS system integration, temperature scaling for calibration improvement, 5-fold cross-validation, and prospective clinical trials comparing CerebAI-assisted reading against radiologist-only base-lines.

## VII. Conclusion

We have presented CerebAI, a clinically motivated system for three-class non-contrast CT stroke classification combining a fine-tuned ConvNeXt-Base backbone with Integrated Gradients attribution. CerebAI achieves 97.47% accuracy, weighted F1 of 0.9746, macro AUC of 0.9921, and Cohen’s *κ* of 0.9483, outperforming ResNet-50, EfficientNet-B4, and ViT-B/16 under identical experimental conditions. The ablation study demonstrates that ImageNet pretraining is essential (F1 −43.43% without it), and the Grad-CAM comparison provides principled justification for choosing Integrated Gradients in medical explainability. Well-calibrated confidence scores (ECE=0.0115), native DICOM support, and pixel-level explanations collectively address the primary obstacles to clinician trust in AI-assisted stroke triage. All code and weights are released to support reproducibility and community advancement.

## Data Availability

The dataset used is publicly available on Kaggle (Orvile, 2024: https://www.kaggle.com/datasets/orvile/inme-veri-seti-stroke-dataset). Code and model weights are available at https://github.com/ar-shenoy. A live demo is hosted at https://huggingface.co/spaces/arshenoy/cerebAI.

https://github.com/ar-shenoy

https://huggingface.co/spaces/arshenoy/cerebAI

## Acknowledgment

The author thanks the open-source contributors of Py-Torch, timm, Captum, and Albumentations. Code and model are available at https://github.com/ar-shenoy and https://huggingface.co/spaces/arshenoy/cerebAI

